# COVID-19 mRNA Booster Vaccines Elicit Strong Protection Against SARS-CoV-2 Omicron Variant in Patients with Cancer

**DOI:** 10.1101/2021.12.28.21268398

**Authors:** Cong Zeng, John P. Evans, Karthik Chakravarthy, Panke Qu, Sarah Reisinger, No Joon Song, Mark P. Rubinstein, Peter G. Shields, Zihai Li, Shan-Lu Liu

## Abstract

Following its emergence in late November of 2020, the SARS-CoV-2 Omicron (B.1.1.529) variant has caused major global public health concerns. We recently demonstrated that in healthy adults the Omicron variant exhibits strong resistance to immunity induced by two doses of the mRNA vaccines, but a booster mRNA vaccine dose can provide strong protection against Omicron. However, it is currently unknown how well these mRNA vaccine boosters protect immunocompromised groups, including cancer patients, from the Omicron variant. Here we show that (1) neutralizing antibody (nAb) titers against the Delta and Omicron variants in cancer patients after two-dose mRNA vaccines are 4.2-fold and 21.3-fold lower, respectively, compared to the ancestral D614G, and (2) nAb titers against the Delta and Omicron variants in boosted cancer patients are 3.6-fold and 5.1-fold lower, respectively, compared to D614G. Our findings highlight the effectiveness and need for booster vaccination strategies in immunocompromised groups including cancer patients to protect from the Omicron variant.

---

The emerging SARS-CoV-2 Omicron (B.1.1.529) variant has caused considerable concern about the future of containing the ongoing COVID-19 pandemic (WHO, 2021). First isolated in late November of 2021, the Omicron variant harbors a staggering 30-40 mutations in the viral spike (S) protein which includes substantial changes to the S receptor-binding domain that produced significant concern about the potentially high immune evasion of this variant (Pulliam et al., 2021). Additionally, the rapid increase in COVID-19 cases attributed to the Omicron variant (Karim et al, 2021) caused a major public health concern about its increased transmissibility. We recently reported that the Omicron variant exhibits drastic resistance to neutralizing antibodies (nAbs) from healthy recipients of two-dose mRNA-1273 or BNT162b2 mRNA vaccines as well as COVID-19 patients (Zeng et al., 2021). However, healthy individuals who received an additional third mRNA booster vaccine dose exhibit much stronger protection against the Omicron variant, comparable to their protection against other SARS-CoV-2 variants of concern (Zeng et al., 2021). Our results indicate that not only does booster vaccination enhances nAb levels, but also broadens the nAb response against these VOCs, including the Omicron.

However, it remains unclear how booster vaccination impacts immunity against the Omicron variant in immunocompromised groups, especially patients with cancer on active therapy. Report from our group has recently shown that patients with cancer are a key group of immunocompromised individuals with reduced responsiveness to two-dose mRNA vaccination (Zeng et al., 2021). This reduced vaccine responsiveness can be somewhat overcome by the administration of booster vaccine doses (Greenberger et al., 2021; Shapiro et al., 2021). However, the breadth of the nAb response in these boosted patients, in particular their immunity against the Omicron variant, remains unclear. This is a critical question to resolve to determine future vaccination strategies, especially the administration of additional booster doses, for immunocompromised groups.

To address this urgent need we utilized our previously reported, highly-sensitive, pseudotyped-lentivirus neutralization assay (Zeng et al., 2020) to examine the nAb response to Omicron compared to Delta and the ancestral D614G variants in patients with cancer (n = 50) following two mRNA vaccine doses (n = 23) and three doses including a third booster vaccine (n = 27). Samples were collected between 31 and 121 days (median 95 days) post-second mRNA vaccine dose for 15 lung cancer patients vaccinated with mRNA-1273 (n = 10) or BNT162b2 (n = 5), and 8 breast cancer patients vaccinated with mRNA-1273 (n = 2) or BNT162b2 (n = 6). For booster vaccine recipients, samples were collected between 2 and 112 days (median 47 days) post-third mRNA vaccine dose for solid tumors, including 12 breast cancer patients vaccinated with mRNA-1273 (n = 4) or BNT162b2 (n = 8) and a mix of 15 other types of solid tumor patients (such as melanoma, genitourinary, gastrointestinal, etc.) vaccinated with either mRNA-1273 (n = 5) or BNT162b2 (n = 10) (**Table S1**).

Neutralizing Ab titers (NT_50_) were measured against the Omicron variant, along with the ancestral D614G SARS-CoV-2 and the Delta variant, the latter being responsible for the most recent wave of infections. We found that for recipients of two mRNA vaccine doses, the Delta and Omicron variants exhibited a 4.2-fold (p < 0.01) and 21.3-fold (p < 0.01) reduction in NT_50_, respectively, compared to D614G (**Fig. S1A**). Notably, 0% (0/23), 13.0% (3/23), and 52.2% (12/23) patients exhibited nAb titers below the limit of detection (NT_50_ < 40, as determined by the lowest dilution used in the neutralization assay) for the D614G, Delta, and Omicron variants (**Fig. S1A)**, respectively. These results clearly demonstrate the strong nAb resistance of the Omicron variant against two mRNA vaccine doses in patients with cancer. Additionally, mRNA-1273 patients exhibited statistically insignificant higher nAb titers compared to those vaccinated with BNT162b2 (**Fig. S1B**).

We further examined the nAb titers against the Omicron, D614G, and Delta variants for patients with cancer who received an additional third dose of vaccination, i.e., a booster dose. Overall, booster recipients exhibited dramatically increased nAb titers (**Fig. S1C**). Strikingly, we found that the Omicron variant only exhibited a 5.1-fold (p < 0.0001) reduction in nAb titers, respectively, compared to D614G (**Fig. S1C**), indicating generation of a stronger and much broader neutralization against the Omicron variant after the booster vaccination. The nAb titer against the Delta variant in booster recipients dropped by 3.6-fold compared to D614G, which was comparable to the two-dose vaccination (**Fig. S1C**). Of note, 0% (0/27), 0% (0/27), and 11.1% (3/27) patients exhibited nAb titers below the limit of detection (NT_50_ < 80; the lowest dilution used was 1:80 because of relatively high titer for boosters) for the D614G, Delta and Omicron variants, respectively (**Fig. S1C**). These results were in sharp contrast to the two-dose vaccination regime, showing that a booster mRNA vaccine dose significantly provided enhanced broader protection against the Omicron variant for patients with cancer, which is similar to the observed protection against Omicron in healthy boosted individuals(1). We found that BNT162b2-boosted patients showed higher nAb titers than mRNA-1273-boosted patients, albeit not statistically significant (**Fig. S1D**).

We also examined the impact of anti-PD-1/PD-L1 treatments on nAb response in two-dose and booster-vaccinated patients with cancer. We observed statistically insignificant NT_50_ titers in non-PD-1/PD-L1-treated patients compared to patients treated with immune checkpoint blockers regardless if they received two-dose or three-dose vaccines including the booster shots (**Fig. S1E**).

Despite previous findings that neutralizing antibody response to mRNA vaccination is age dependent (Collier et al., 2021), we did not observe a statistically significant correlation between age and NT_50_ values in these patients with cancer (**Figs. S1F**), which was likely due to the older populations and the relatively small size of this cohort. Given increasing concerns about declining efficacy of SARS-CoV-2 vaccines, especially against variants of concern (VOCs) (Keehner et al., 2021), we also examined the correlation between NT_50_ and time post second vaccine dose for these patients with cancer. Indeed, we observed a negative correlation between time after second and booster doses of mRNA vaccination and NT_50_ value (**Figs. S1G**); however, statistically they were not significant, again likely because of the limited sample sizes in this cohort. Nevertheless, these results suggested possible waning immune protection of neutralizing antibodies against SARS-CoV-2 and VOCs following mRNA vaccination, including the booster.

We investigated the efficacy of mRNA vaccine-induced neutralizing antibody response in patients with solid tumors against the Omicron variant, in parallel with the ancestral D614G and the Delta variants. Our results demonstrate that patients with cancer who received booster dose of mRNA vaccine displayed a significantly greater neutralizing capacity against the Omicron variant in comparison to those recipients of two-doses of mRNA vaccine. This result aligns with findings in health care workers (HCWs) (Zeng et al., 2021). While the mechanism underlying the enhanced breadth of the neutralizing antibody activity against Omicron, and likely other emerging variants, requires further investigation, our results provide reassurance that a booster after two-dose mRNA vaccination enhances protection in patients with solid tumors.

The recent emergence of variants of concern for SARS-CoV-2 has made the impact of vaccine response in immunocompromised individuals, particularly patients with cancer, a topic of significant interest. Our study demonstrated that the diagnosis of a solid cancer *per se* does not appear to negatively impact the adaptive immune response to booster-mediated protections against SARS-CoV-2 variants, including Omicron. Some of the limitations of this study include the small size of the cohort (n = 50 patients with cancer), the differences between the two-dose and booster cohorts regarding solid tumor subtypes, as well as timing of sample collection post vaccination. Additionally, the utilization of a single assay to evaluate nAb levels may not be fully representative of the status of infection protection in these patients with cancer. Finally, our study did not address the risk of breakthrough infections clinically. Therefore, patients with cancer should still remain cautious regarding immunity to the Omicron variant even after vaccination with an mRNA booster.

## Data Availability

All data produced in the present study are available upon reasonable request to the authors.

## Acknowledgements

We thank members of the Liu lab for sharing reagents and discussion. Additonal gratitude is extended towards Kevin Weller, Jamie Hamon, Donna Bucci, Kelsi Reynolds, Mohamed Yusuf, Robert Davenport, and Taylor Chatlos from the Immune Monitoring and Discovery Platform of the Pelotonia Institute for Immuno-Oncology for assistance in sample collection and discussion. We also thank members of the Li lab, including Chelsea Bolyard, Weiwei Liu, and Yi Wang, and members of the Rubinstein Lab, including Madison Sikorski, Joseph Azar, Rachael Teodorescu, Ian Carmody, Selah McKenney, and Kayla Hudson, for assistance in sample processing. Finally, we would like to thank the NIH AIDS Reagent Program and BEI Resources for supplying important reagents that made this work possible. This work was supported by a fund provided by an anonymous private donor to The Ohio State University to S.-L.L. S.-L.L. was additionally supported in part by NIH R01 AI150473. Z.L. was supported by The Ohio State University Comprehensive Cancer Center Cancer Center Support Grant (P30CA016058), and multiple NIH grants (R01 DK105033; R01 AI077283; R01 CA213290; P01 CA186866; R01 CA255334; P30 CA016058). P.G.S. was supported by multiple NIH grants including NCI P30 CA016058. S.R. and Z.L. were supported by the Peletonia Institute for Immune-Oncology. The Pelotonia Institute for Immuno-Oncology (PIIO) is funded in part by the Pelotonia community and the Ohio State University Comprehensive Cancer Center (OSUCCC) (P30CA016058). The support from the OSUCCC Shared Resources includng Flow Cytometry and Clinical Treatment Unit and Clinical Trials Processing Laboratory is greatly appreciated.

## Supplementary Methods

### Patient Cohorts

Sera were obtained from patients with cancer under an approved IRB protocol (2021C0041). Samples were collected between 31 and 121 days post-second mRNA vaccine dose (n = 23) for 8 breast cancer patients (all female) vaccinated with mRNA-1273 (n = 2) or BNT162b2 (n = 6) and for 15 lung cancer patients (6 female and 9 male) vaccinated with mRNA-1273 (n = 10) or BNT162b2 (n = 5). These patients had a median age of 63 (IQR = 13).

For booster vaccine recipients (n = 27), samples were collected between 2 and 112 days post second mRNA vaccine dose for 12 breast cancer patients (all females) vaccinated with mRNA-1273 (n = 4) or BNT162b2 (n = 8); 5 genitourinary cancer, 5 gastrointestinal cancer, 3 melanoma, 1 sarcoma, and 1 thymic cancer patients vaccinated with mRNA-1273 (n = 5) or BNT162b2 (n = 10); these patients had a median age of 58 (IQR = 13).

### Plasmids

For pseudotyped virus production we utilized our previously reported pNL4-3-inGluc lentivirus vector (Zeng, et al., 2020). This vector is based on ΔEnv HIV-1 and bears a *Gaussia* luciferase reporter gene in anti-sense orientation relative to the HIV-1 genome. The reporter gene then contains an intron oriented in the sense orientation relative to the HIV-1 genome to prevent *Gaussia* luciferase expression in the virus producer cells. Additionally, we utilized N-and C-terminally Flag-tagged SARS-CoV-2 S expression constructs that were generated and cloned into pcDNA3.1 using Kpn I and BamH I restriction enzyme cloning by GenScript Biotech (Piscataway, NJ). The constructs contained the following mutations, D614G: D614G; Delta (B.1.617.2): T19R, Δ156-158, L452R, T478K, D614G, P681R, D950N; and Omicron (B.1.1.529): A67V, Δ69-70, T95I, G142D, Δ143-145, N211I, L212VPE, V213P, R214E, G339D, R346K, S371L, S373P, S375F, K417N, N440K, G446S, S477N, T478K, E484A, Q493R, G496S, Q498R, N501Y, Y505H, T547K, D614G, H655Y, N679K, P681H, N764K, D796Y, N856K, Q954H, N969K, and L981F.

### Cell Lines and Maintenance

HEK293T (ATCC CRL-11268, CVCL_1926) and HEK293T-ACE2 (BEI NR-52511) cells were maintained in DMEM (Gibco, 11965-092) supplemented with 10% FBS (Sigma, F1051) and 1% penicillin-streptomycin (HyClone, SV30010) and kept at 37°C and 5% CO_2_.

### Virus Production and Titering

Lentiviral pseudotypes were produced as previously reported (Zeng, et al., 2020; Zeng, et al, 2021). HEK293T cells were transfected with pNL4-3-inGluc and various S constructs in a 2:1 ratio using polyethylenimine transfection. Virus was collected 24, 48, and 72 hrs after transfection, pooled, and stored at −80°C. To determine the relative infectivity of pseudotypes, virus was used to infect HEK293T-ACE2 cells and media from infected cells was assayed for *Gaussia* luciferase activity 48 and 72 hrs after infection, by combining 20 μL of cell culture media with 20 μL of *Gaussia* luciferase substrate (0.1 M Tris (MilliporeSigma, #T6066) pH 7.4, 0.3 M sodium ascorbate (Spectrum, S1349), 10 μM coelenterazine (GoldBio, CZ2.5)). Luminescence measurements were taken with a BioTek Cytation5 plate reader.

### Pseudotyped Lentivirus Neutralization Assays

Pseudotyped lentivirus neutralization assays were performed as previously described (Zeng, et al., 2020; Zeng, et al, 2021). Briefly, patient sera were 4-fold serially diluted and then incubated with pseudotyped virus at 37°C for 1 hr (final dilutions of 1:40, 1:160, 1:640, 1:2560, 1:10240, and no serum control). Following incubation, sera and virus was transferred to seeded HEK293T-ACE2 cells for infection. Media of infected cells was assayed for *Gaussia* luciferase activity 48 and 72 hrs after infection by combining 20 μL of cell culture media with 20 μL or *Gaussia* luciferase substrate. Luminescence was measured by a BioTek Cytation5 plate reader. 50% neutralization titers (NT_50_) values were calculated from luminescence readout by least-squares-fit, non-linear regression performed in GraphPad Prism 5 (San Diego, CA).

### Statistics

Comparisons between multiple groups were made using one-way ANOVA with Bonferroni’s multiple testing correction. Comparisons between two groups were made using two-tailed t-test with Welch’s correction.

## Supplemental Figure and figure legends

**Figure S1.**
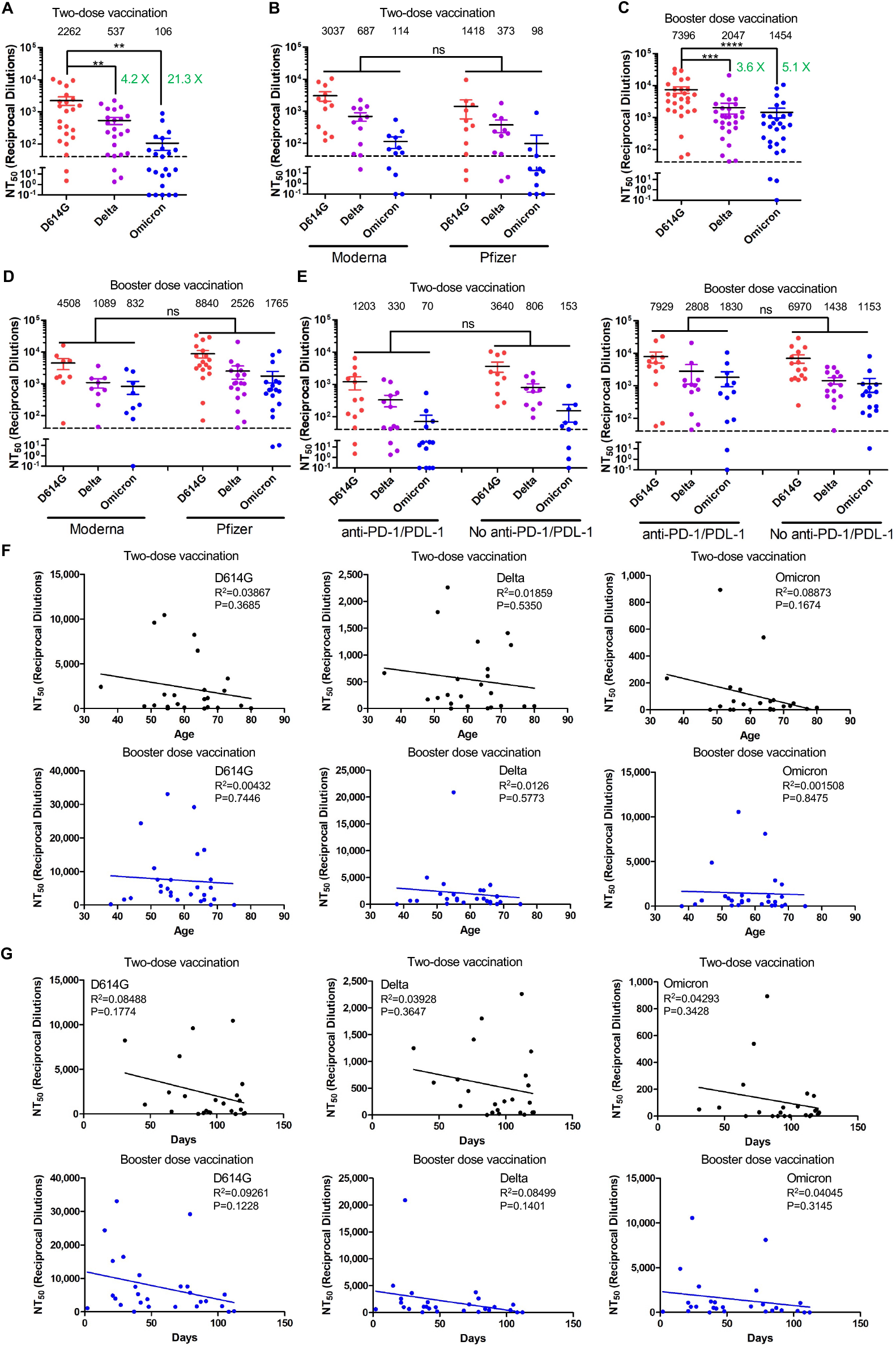
The Omicron variant exhibits strong immune-escape in two-dose-vaccinated patients with cancer which is overcome by booster vaccinations. *Gaussia* luciferase reporter gene-bearing lentiviral vector was pseudotyped with spike from SARS-CoV-2 variant of interest, and pseudotyped viruses were then incubated with serially-diluted patient serum before being used to infect HEK293T-ACE2 cells. Culture media were changed after overnight infection and were measured for the luciferase activity at 48 and 72 hrs post-infection. (**A** and **B**) Sera from 23 patients with solid cancer collected after second mRNA vaccine dose (12 mRNA-1273 and 11 BNT16b2) were used to neutralize pseudotyped virus bearing the spike of D614G, Delta and Omicron variants, and resulting 50% neutralization titers (NT_50_) were calculated. (**C** and **D**) Sera from 27 patients with solid cancer collected after booster vaccination (9 mRNA-1273 and 18 BNT16b2) were used to neutralize pseudotyped viruses of D614G, Delta or Omicron variants, with NT_50_ calculated. (**E**) NT_50_ values against D614G, Delta and Omicron variants in cancer patients, with or without anti-PD-1/PDL-1 treatments: two-dose vaccination and booster dose vaccination. (**F**) Correlative analyses between NT_50_ values against variants and patient’s age: two-dose vaccination and booster vaccination. (**G**) Correlative analyses between NT_50_ against variants and sera collection days: two-dose vaccination and booster vaccination. In all cases, mean NT_50_ values are displayed at the top of each plot; bars represent mean +/-standard error, and significance is determined by one-way ANOVA with Bonferroni’s multiple testing correction. P-values are represented as ** p < 0.01, *** p < 0.001, **** p < 0.0001; ns, not significant (defined as P > 0.05).

**Table S1.** Demographic information of patients with cancer.

|  |  | Total (n=50) |  | Two-Dose (n=23) |  | Booster (n=27) |  |
| --- | --- | --- | --- | --- | --- | --- | --- |
|  |  | n | (%) | n | (%) | n | (%) |
| Age Group (years) | 30-44 | 4 | 8.0% | 1 | 4.3% | 3 | 11.1% |
|  | 45-59 | 20 | 40.0% | 9 | 39.1% | 11 | 40.7% |
|  | 60-74 | 23 | 46.0% | 11 | 47.9% | 12 | 44.5% |
|  | 75-85 | 3 | 6.0% | 2 | 8.7% | 1 | 3.7% |
| Sex | Male | 16 | 32.0% | 9 | 39.1% | 7 | 25.9 |
|  | Female | 34 | 68.0% | 14 | 60.9% | 20 | 74.1 |
| Race | African American/Black | 3 | 6.0% | 2 | 8.7% | 1 | 3.7% |
|  | Asian Chinese | 1 | 2.0% | 0 | 0.0% | 1 | 3.7% |
|  | White | 45 | 90.0% | 21 | 91.3% | 24 | 88.9% |
|  | American Indian/Alaskan | 1 | 2.0% | 0 | 0.0% | 1 | 3.7% |
| Vaccine | mRNA-1273 | 21 | 42.0% | 12 | 52.2% | 9 | 33.3% |
|  | BNT162b2 | 29 | 58.0% | 11 | 47.8% | 18 | 66.7% |
| Solid Tumor Type | Lung | 15 | 30.0% | 15 | 65.2% | 0 | 0.0% |
|  | Breast | 20 | 40.0% | 8 | 34.8% | 12 | 44.4% |
|  | Gastrointestinal | 5 | 10.0% | 0 | 0.0% | 5 | 18.5% |
|  | Genitourinary | 5 | 10.0% | 0 | 0.0% | 5 | 18.5% |
|  | Melanoma | 3 | 6.0% | 0 | 0.0% | 3 | 11.2% |
|  | Thymic carcinoma | 1 | 2.0% | 0 | 0.0% | 1 | 3.7% |
|  | Sarcoma | 1 | 2.0% | 0 | 0.0% | 1 | 3.7% |
| WBC (k/μL) | <4.5 | 13 | 26.0% | 4 | 17.4% | 9 | 33.3% |
|  | 4.5–11.0 | 32 | 64.0% | 15 | 65.2% | 17 | 63.0% |
|  | >11.0 | 5 | 10.0% | 4 | 17.4% | 1 | 3.7% |
| Lymphocyte Count (k/μL) | <1.0 | 20 | 40.0% | 12 | 52.2% | 8 | 3.0% |
|  | 1.0–4.8 | 29 | 58.0% | 10 | 43.5% | 19 | 70.0% |
|  | >4.8 | 1 | 2.0% | 1 | 4.3% | 0 | 0.0% |
| Immunotherapy | Anti-PD-1/PD-L1* | 25 | 50.0% | 13 | 56.5% | 12 | 44.4% |

\*The anti-PD-1/PD-L1 drugs include Nivolumab, Pembrolizumab, Durvalumab, Atezolizumab, and Avelumab.

## Notes

### Competing Interest Statement

The authors have declared no competing interest.

### Author Declarations

This work was performed under an approved IRB protocol #2021C0041 by The Ohio State University Biomedical Sciences Institutional Review Board.

## References

Collier, D.A., Ferreira, I.A., Kotagiri, P., Datir, R.P., Lim, E.Y., Touizer, E., Meng, B., Abdullahi, A., Elmer, A., and Kingston, N., et al. (2021). Age-related immune response heterogeneity to SARS-CoV-2 vaccine BNT162b2. Nature 596, 417–422.

Greenberger, L.M., Saltzman, L.A., Senefeld, J.W., Johnson, P.W., DeGennaro, L.J., and Nichols, G.L. (2021). Anti-spike antibody response to SARS-CoV-2 booster vaccination in patients with B cell-derived hematologic malignancies. Cancer Cell.

Karim, S.S.A., and Karim, Q.A. (2021). Omicron SARS-CoV-2 variant: a new chapter in the COVID-19 pandemic. The Lancet.

Keehner, J., Horton, L.E., Binkin, N.J., Laurent, L.C., Pride, D., Longhurst, C.A., Abeles, S.R., and Torriani, F.J. (2021). Resurgence of SARS-CoV-2 infection in a highly vaccinated health system workforce. New England Journal of Medicine 385, 1330–1332.

Pulliam, J.R.C., Schalkwyk, C.V., Govender, N., Gottberg, A.V., Cohen, C., Groome, M., Dushoff, J., Mlisana, K., and Moultrie, H. (2021). Increased risk of SARS-CoV-2 reinfection associated with emergence of the Omicron variant in South Africa. MedRxiv.

Shapiro, L.C., Thakkar, A., Campbell, S.T., Forest, S.K., Pradhan, K., Gonzalez-Lugo, J.D., Quinn, R., Bhagat, T.D., Choudhary, G.S., and McCort, M., et al. (2021). Efficacy of booster doses in augmenting waning immune responses to COVID-19 vaccine in patients with cancer. Cancer Cell.

World Health Organization (2021). Classification of Omicron (B.1.1.529):SARS-CoV-2 Variant of Concern.

Zeng, C., Evans, J.P., Pearson, R., Qu, P., Zheng, Y.-M., Robinson, R.T., Hall-Stoodley, L., Yount, J., Pannu, S., and Mallampalli, R.K., et al. (2020). Neutralizing antibody against SARS-CoV-2 spike in COVID-19 patients, health care workers, and convalescent plasma donors. JCI insight 5.

Zeng, C., Evans, J.P., Qu, P., Faraone, J.N., Zheng, Y.-M., Carlin, C., Bednash, J.S., Zhou, T., Lozanski, G., Mallampalli, R., et al. (2021a). Neutralization and stability of SARS-CoV-2 Omicron variant. BioRxiv.

Zeng, C., Evans, J.P., Reisinger, S., Woyach, J., Liscynesky, C., Boghdadly, Z.E., Rubinstein, M.P., Chakravarthy, K., Saif, L., and Oltz, E.M., et al. (2021b). Impaired neutralizing antibody response to COVID-19 mRNA vaccines in cancer patients. Cell & Bioscience 11, 1–6.

## Supplemental References

Zeng, C., Evans, J.P., Qu, P., Faraone, J.N., Zheng, Y.-M., Carlin, C., Bednash, J.S., Zhou, T., Lozanski, G., Mallampalli, R., et al. (2021). Neutralization and stability of SARS-CoV-2 Omicron variant. BioRxiv.

